# Effect of Covid-19 on inequalities in premature mortality in England: an analysis of excess mortality by deprivation and ethnicity

**DOI:** 10.1101/2021.05.18.21256717

**Authors:** Sharmani Barnard, Paul Fryers, Justine Fitzpatrick, Sebastian Fox, Allan Baker, Paul Burton, John Newton, Yvonne Doyle, Peter Goldblatt

## Abstract

**Objectives:** To examine the impact of the Covid-19 pandemic on inequalities in premature mortality in England by deprivation and ethnicity.

**Design:** A statistical model to estimate increased mortality in population sub-groups during the Covid-19 pandemic by comparing observed with expected mortality in each group based on trends over the previous five years.

**Setting:** Information on deaths registered in England since 2015 was used, including age, sex, area of residence, and cause of death. Ethnicity was obtained from Hospital Episode Statistics (HES) records linked to death registration data.

**Participants:** Population study of England, including all 569,824 deaths from all causes registered between 21 March 2020 and 26 February 2021.

**Main outcome measures:** Excess mortality in each sub-group over and above the number expected based on trends in mortality in that group over the previous five years.

**Results:** The gradient in excess mortality by deprivation was greater in the under 75s (most deprived had 1.25 times as many deaths as expected, least deprived 1.14) than in all ages (most deprived had 1.24 times as many deaths as expected, least deprived 1.20). Among the Black and Asian groups, all deprivation quintiles had significantly larger excesses than the most deprived White group and there were no clear gradients across quintiles. Among the White group, only the most deprived had more excess deaths than deaths directly involving Covid-19. Among the Black group all deprivation quintiles experienced more excess deaths than deaths directly involving Covid-19.

**Conclusion:** The Covid-19 pandemic has widened inequalities in premature mortality by deprivation. Among those under 75, the direct and indirect effects of the pandemic on deaths have disproportionately impacted ethnic minority groups irrespective of deprivation, and the most deprived White group. Statistics limited to deaths directly involving Covid-19 understate the pandemic’s impact on inequalities by deprivation and ethnic group at younger ages.

**Key Messages:** 

**What is already known on this topic:**

- Estimates of excess mortality for all ages combined for the period March 2020 to February 2021 show a small difference in the relative impact of Covid-19 on excess mortality between the most deprived and the least deprived
- Throughout the pandemic estimates of excess mortality have been greater for ethnic minority groups compared with the white group.

**What this study adds:**

- Among the under 75s the gradient of excess mortality across the deprivation spectrum indicates a stark increase in already established inequalities in premature mortality
- Independent of deprivation, excess mortality in Black and Asian ethnic groups is much higher than that of the white group, indicating that ethnicity is a determinant of excess mortality, regardless of level of area-based deprivation.

## Introduction

Inequalities in mortality from Covid-19 have been widely reported by age, gender, deprivation and ethnic group (1-5). Mortality from Covid-19 has been highest among older groups, with people aged over 80 years estimated to have 70 times the risk of mortality from Covid-19 once infected, compared with those aged under 40 years (3). Mortality among males has been higher than in females and after controlling for age and co-morbidities, mortality among many ethnic minority groups has been higher than the White group (3, 6, 7).

There is consistent evidence that mortality rates for deaths associated with Covid-19 during the first wave of the pandemic were more than twice as high among the most deprived as in the least deprived (3, 5, 8). However, all-cause mortality was also higher among the most deprived groups in previous years (3, 9).

Measures of excess mortality compare the mortality that would be expected in each group, based on trends in previous years, with what has occurred in that group (4, 10, 11). They therefore take account of the existing inequalities in the baseline and show the extent to which Covid-19 has impacted on inequalities between groups. Estimates of excess mortality for all ages combined for the period March 2020 to February 2021 show a small difference in the relative impact of Covid-19 on excess mortality between the most deprived and the least deprived, and show a greater excess in ethnic minority groups (12). However, these estimates have not been presented separately for each ethnic group by deprivation. In addition, these overall estimates are heavily weighted towards deaths at older ages because of the far higher death rates in older people.

Understanding the relationship between deprivation, ethnicity and excess mortality among the those aged under 75 years is important for four reasons. First, there is an existing gradient in mortality among deprivation groups; in the years prior to the Covid-19 pandemic a third of premature deaths (<75 years) were attributable to socioeconomic inequality (13). Second, when comparing the mortality that has occurred during the pandemic with what would have been expected in the absence of the pandemic, the relative increase has been highest among those aged 45-64 years (12). Third, we know that Covid-19 death rates have been higher in ethnic minority groups and deprived areas, but we do not fully understand the interaction between the two (3). Fourth, the deprivation quintile allocated to care homes based on their address, where a significant proportion of people over 75 reside, may not accurately reflect the lifetime deprivation of the care home residents and may play a role in masking the relationship between excess mortality and deprivation at older ages.

In this study, we investigate the relationship between age, deprivation, ethnicity and mortality, by estimating excess mortality for each deprivation quintile among those aged under 75 (premature mortality) for each ethnic group. By doing this we aim to identify the relative effect of the Covid-19 pandemic on mortality for each ethnic group in each deprivation quintile, based on what would have been expected for these groups. This will therefore identify any increases in inequalities that are over and above existing inequalities in mortality between deprivation quintiles in each ethnic group.

## Methods

### Setting

The first reported death due to Covid-19 in England was on 5 March 2020 (14). PHE began receiving a daily feed of registered deaths from the Office for National Statistics (ONS) on 21 March 2020 to enable timely analysis of mortality during the pandemic. We examined excess mortality in England by deprivation quintile from 21 March 2020 to 26 February 2021. All registered deaths of usual residents of England in this period (N=569,824) were included in the analysis. All those deaths recorded as mentioning Covid-19 on the death certificate (directly involving Covid-19) in England until 26 February 2021 were thus incorporated in the analysis, except for 76 Covid-19 deaths registered before 21 March 2020. All analyses were carried out on 10 March 2021.

### Measuring excess mortality

#### Analyses

All analyses included deaths from all causes. We investigated excess mortality by deprivation quintile separately among all persons of all ages and among those aged under 75 years. We also present excess mortality estimates for each deprivation quintile by ethnic group among those aged under 75.

#### Data and Methods

The number of excess deaths in each group is the number of observed deaths relative to the number of deaths that would have been expected based on trends in data from the previous five years for that group (baseline deaths). The model used to estimate expected deaths accounts for trends in mortality and demographic changes. The data and methods used by PHE to estimate excess mortality for weekly national excess reports were adopted for this analysis (12). Deprivation is measured by the Index of Multiple Deprivation(15) for the Lower Super Output Area (LSOA) of residence. LSOAs are allocated to national quintiles based on the deprivation score. Ethnicity is measured at the individual level through linkage to Hospital Episode Statistics.

Further details of the data sources and statistical modelling are available in the methodology documentation included with the weekly reports (11). All data were analysed using the generalised linear modelling function in the statistical package R (version 4.0.3) (16).

## Results

### Excess mortality by deprivation quintile among all persons

Between 21 March 2020 and 26 February 2021, the model estimates there were 101,501 excess deaths among all persons in England. This is 1.22 times the number of deaths expected, had there been no pandemic. There was a slight gradient in relative excess mortality from the most deprived quintile of the population (1.24 times the expected; 99.8% CI 1.24–1.25) to the least deprived (1.20 times; 99.8% CI 1.19–1.21) (Figure 1). In every deprivation quintile, the number of deaths with Covid-19 mentioned on the death certificate (124,437 in all) exceeded the number of excess deaths (Table 1).

**Table 1:** Excess mortality by deprivation quintile among all persons, 21 March 2020 to 26 February 2021

| Deprivation Quintile | Registered deaths | Expected deaths | Ratio registered / expected | Excess deaths | Upper CL | Lower CL | Covid-19 deaths |
| --- | --- | --- | --- | --- | --- | --- | --- |
| **Quintile 1 - Most Deprived** | 120,462 | 96,808 | 1.24 | 23,654 | 1.24 | 1.25 | 28,403 |
| **Quintile 2** | 114,821 | 93,918 | 1.22 | 20,903 | 1.21 | 1.23 | 26,507 |
| **Quintile 3** | 116,403 | 96,056 | 1.21 | 20,347 | 1.20 | 1.22 | 24,524 |
| **Quintile 4** | 113,002 | 94,171 | 1.20 | 18,831 | 1.19 | 1.21 | 23,529 |
| **Quintile 5 - Least Deprived** | 105,136 | 87,370 | 1.20 | 17,766 | 1.19 | 1.21 | 21,474 |
| **Total** | 569,824 | 468,323 | 1.22 | 101,501 | 1.21 | 1.22 | 124,437 |

### Excess mortality by deprivation quintile among persons aged <75

Among those aged under 75 years, 177,809 deaths were registered in the same period – 29,845 more than expected or 1.20 times (99.8% CI 1.19–1.21) the number expected based on trends in previous years (Table 2). There was a steeper gradient in excess mortality by deprivation quintile compared with the all ages analysis, with excess mortality highest among the most deprived (1.25 times the expected; 99.8% CI 1.24–1.27) and lowest among the least deprived (1.14 times; 99.8% CI 1.12–1.16) (Figure 2). This difference in gradient largely results from lower excess mortality in the three least deprived quintiles among those under 75 years, compared with all age groups. Among the most deprived quintile, there were more excess deaths than deaths with Covid-19 on the death certificate (Table 2).

**Table 2:**
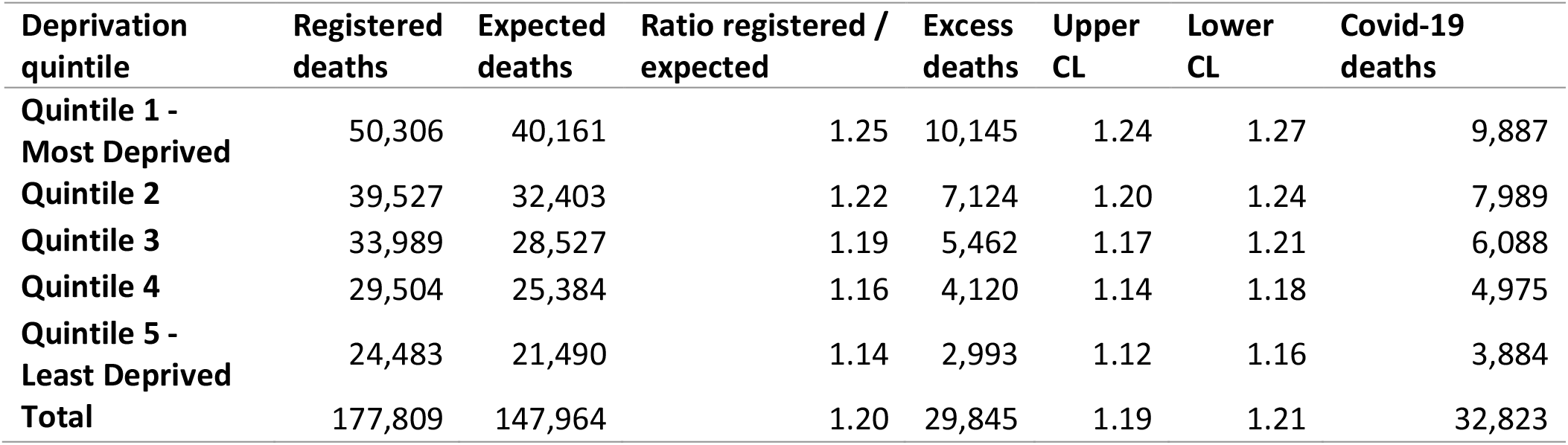
Excess mortality by deprivation quintile among people aged <75 years 21 March 2020 to 26 February 2021

| Deprivation quintile | Registered deaths | Expected deaths | Ratio registered / expected | Excess deaths | Upper CL | Lower CL | Covid-19 deaths |
| --- | --- | --- | --- | --- | --- | --- | --- |
| **Quintile 1 - Most Deprived** | 50,306 | 40,161 | 1.25 | 10,145 | 1.24 | 1.27 | 9,887 |
| **Quintile 2** | 39,527 | 32,403 | 1.22 | 7,124 | 1.20 | 1.24 | 7,989 |
| **Quintile 3** | 33,989 | 28,527 | 1.19 | 5,462 | 1.17 | 1.21 | 6,088 |
| **Quintile 4** | 29,504 | 25,384 | 1.16 | 4,120 | 1.14 | 1.18 | 4,975 |
| **Quintile 5 - Least Deprived** | 24,483 | 21,490 | 1.14 | 2,993 | 1.12 | 1.16 | 3,884 |
| **Total** | 177,809 | 147,964 | 1.20 | 29,845 | 1.19 | 1.21 | 32,823 |

### Deprivation by ethnic group among the <75s

Excess mortality among those aged under 75 years by ethnic group and deprivation quintile is presented in Figure 3 and Table 3. Among those under 75, the White population saw the smallest relative excess compared with the number expected (1.17; 99.8% CI 1.16–1.17). The Asian (1.63; 99.8% CI 1.59–1.67) and Black (1.58; 99.8% CI 1.53–1.63) populations saw the largest relative increase in deaths compared with the number of deaths expected based on previous years.

**Table 3:** Excess mortality by deprivation quintile and ethnic group among people aged <75 years 21 March 2020 to 26 February 2021

| **Ethnic group and deprivation quintile** | **Registered deaths** | **Expected deaths** | **Ratio registered / expected** | **Excess deaths** | **Upper CL** | **Lower CL** | **Covid-19 deaths** |
| --- | --- | --- | --- | --- | --- | --- | --- |
| **Asian Group** |  |  |  |  |  |  |  |
| Quintile 1 - Most Deprived | 3,590 | 2,284 | 1.57 | 1,306 | 1.51 | 1.64 | 1,475 |
| Quintile 2 | 2,959 | 1,728 | 1.71 | 1,231 | 1.64 | 1.78 | 1,261 |
| Quintile 3 | 1,782 | 1,078 | 1.65 | 704 | 1.57 | 1.74 | 700 |
| Quintile 4 | 1,145 | 701 | 1.63 | 444 | 1.52 | 1.74 | 422 |
| Quintile 5 - Least Deprived | 808 | 506 | 1.60 | 302 | 1.47 | 1.74 | 271 |
| Total Asian | 10,285 | 6,297 | 1.63 | 3,988 | 1.59 | 1.67 | 4,129 |
| **Black Group** |  |  |  |  |  |  |  |
| Quintile 1 | 2,240 | 1,487 | 1.51 | 753 | 1.43 | 1.59 | 701 |
| Quintile 2 | 1,851 | 1,143 | 1.62 | 707 | 1.53 | 1.71 | 609 |
| Quintile 3 | 809 | 503 | 1.61 | 306 | 1.48 | 1.73 | 260 |
| Quintile 4 | 428 | 246 | 1.74 | 182 | 1.57 | 1.93 | 129 |
| Quintile 5 | 219 | 135 | 1.62 | 84 | 1.38 | 1.90 | 54 |
| Total Black | 5,547 | 3,515 | 1.58 | 2,032 | 1.53 | 1.63 | 1,754 |
| **Mixed Group** |  |  |  |  |  |  |  |
| Quintile 1 | 507 | 329 | 1.54 | 178 | 1.38 | 1.71 | 110 |
| Quintile 2 | 322 | 253 | 1.27 | 69 | 1.07 | 1.46 | 82 |
| Quintile 3 | 255 | 185 | 1.38 | 70 | 1.17 | 1.60 | 64 |
| Quintile 4 | 163 | 144 | 1.13 | 19 | 0.90 | 1.39 | 42 |
| Quintile 5 | 119 | 107 | 1.11 | 12 | 0.84 | 1.41 | 28 |
| Total Mixed | 1,365 | 1,018 | 1.34 | 347 | 1.25 | 1.43 | 327 |
| **Other Group** |  |  |  |  |  |  |  |
| Quintile 1 | 1,025 | 660 | 1.55 | 365 | 1.44 | 1.67 | 291 |
| Quintile 2 | 1,080 | 661 | 1.63 | 418 | 1.53 | 1.75 | 364 |
| Quintile 3 | 694 | 453 | 1.53 | 241 | 1.39 | 1.66 | 203 |
| Quintile 4 | 449 | 364 | 1.24 | 86 | 1.08 | 1.39 | 113 |
| Quintile 5 | 382 | 298 | 1.28 | 84 | 1.12 | 1.46 | 82 |
| Total Other | 3,630 | 2,436 | 1.49 | 1,194 | 1.43 | 1.55 | 1,054 |
| **White Group** |  |  |  |  |  |  |  |
| Quintile 1 | 42,944 | 35,401 | 1.21 | 7,544 | 1.20 | 1.23 | 7,309 |
| Quintile 2 | 33,316 | 28,618 | 1.16 | 4,698 | 1.15 | 1.18 | 5,672 |
| Quintile 3 | 30,449 | 26,308 | 1.16 | 4,140 | 1.14 | 1.18 | 4,861 |
| Quintile 4 | 27,318 | 23,928 | 1.14 | 3,390 | 1.12 | 1.16 | 4,269 |
| Quintile 5 | 22,955 | 20,444 | 1.12 | 2,511 | 1.10 | 1.14 | 3,450 |
| Total White | 156,983 | 134,699 | 1.17 | 22,283 | 1.16 | 1.17 | 25,561 |

Among the White group, there was a clear gradient of relative excess between the most deprived (1.21; 99.8% CI 1.20–1.23) and the least deprived (1.12; 99.8% CI 1.10–1.14). In the Asian population, all deprivation groups experienced a relative excess between 1.57 (99.8% CI 1.51–1.64) and 1.71 times (99.8% CI 1.64–1.78) but there was no obvious gradient with deprivation, and no significant differences between the quintiles. In the Black population, all deprivation groups had between 1.51 (99.8% CI 1.43–1.59) and 1.74 times (99.8% CI 1.57–1.93) the expected deaths, again with no clear gradient and no significant differences between the quintiles. However, among the Black and Asian groups, all deprivation quintiles saw far greater excesses; significantly greater than even the most deprived in the White group. In the Mixed and Other groups there is the suggestion of a gradient, with higher excess in the more deprived groups, but with very large confidence intervals.

Excess deaths exceeded deaths directly involving Covid-19 in some quintiles in all ethnic groups, although the deprivation quintiles where this occurred differed between ethnic groups (Table 3). In the White ethnic group, an excess greater than the number involving Covid-19 was experienced only in the most deprived group. In contrast, among the Black group all deprivation quintiles experienced excess deaths greater than those directly involving Covid-19. Among the Asian group excess deaths exceeded Covid-19 deaths by a small amount in the three least deprived groups. Full details of excess mortality by deprivation group for ethnic groups are presented in Table 3.

## Discussion

Among those aged under 75 years, excess mortality (compared with expected deaths based on that group’s mortality trends in 2015–2019) was clearly associated with deprivation: highest among the most deprived and lowest among the least deprived. Since higher mortality rates among the more deprived groups prior to the pandemic are taken into account in modelling the expected deaths, this gradient across the deprivation spectrum indicates a stark increase in already established inequalities in premature mortality.

Excess mortality was also clearly associated with ethnicity, with higher overall excess in the Black, Asian, Mixed and Other ethnic groups than the White group. However, the relationship between excess mortality and deprivation is not consistent across ethnic groups: while the White group saw a clear positive gradient between excess mortality and increased deprivation, among Black and Asian groups there was no clear gradient and all deprivation quintiles saw far greater excess than even the most deprived White group. This indicates that ethnicity is a determinant of excess mortality, regardless of level of area-based deprivation.

We also provide evidence that the extent to which excess deaths are directly attributable to Covid-19 is not consistent between ethnic groups and across deprivation quintiles. The Black and Other groups showed clear evidence of excess deaths over and above deaths directly involving Covid-19 across the range of deprivation. In the Asian group the excess deaths above the numbers of deaths involving Covid-19 were small and only in the three least deprived groups. In the White group, it was only the most deprived group that saw excess deaths greater than deaths involving Covid-19.

These excess deaths not attributable to Covid-19 in the White most deprived group and more generally among ethnic minority groups may indicate a lack of recording of Covid-19 on death certificates when testing was limited at the start of the pandemic, or deaths indirectly caused by the pandemic. This may be through factors such as more limited access to healthcare for conditions other than Covid-19, the impact of measures taken to combat the spread of the virus or other consequences of the pandemic. This warrants further analysis and investigation; it is possible that the impact could be reduced by ensuring access to the NHS, take up of Covid-19 testing, and addressing the wider impact of lockdown, such as mental health and financial issues. Understanding how these factors affect ethnic groups differentially is critical to reducing inequalities.

The prevalence of many long-term health conditions is greater in some ethnic minority populations and deprived communities which may also be a factor (17-19). One survey, based on data up to the end of January 2021, found 51.9% of the of the population did not seek help for a worsening health condition during the pandemic (20). Long-term conditions have very different treatment pathways and the wider impact of the pandemic may not be the same for all. For example, hypertensive symptoms or increasing angina are signals for attendance at A&E, but a growing or deteriorating cancer would have been affected by the cancellation of elective clinics, referrals from primary care, or postponement of screening services. These differential impacts of this may not yet be fully realised or understood.

This study shows a greater widening of inequality in premature mortality than deaths at all ages. However, the impact of the pandemic on people in care homes has been substantial and 24% of all deaths involving Covid-19 between 21 March 2020 and 26 February 2021 occurred in care homes (12). The large number of people aged over 75 years in care home facilities, where the deprivation quintile of the care home may not accurately reflect the lifetime deprivation of the care home residents, may play a role in altering the relationship between excess mortality and deprivation at older ages. In addition, the Vivaldi study has shown that there are many other factors that have influenced the impact of the pandemic on individual care homes such as location, characteristics of the home and staffing arrangements which may mean this relationship is not straightforward (21).

This study has many strengths. It is the first study to investigate the relationships between deprivation and excess mortality during the pandemic among those aged less than 75 and among ethnic groups in England, and to identify important differences across sub-groups. The method we employed provides a useful tool to identify inequalities in excess mortality among different groups because baseline deaths are constructed on what would be expected, given the trends and demographic structures of the underlying populations in each group. It estimates the overall net effect of the pandemic, including both deaths involving Covid-19 and deaths caused by the wider impact of the pandemic.

However, this study does have some limitations. The ethnic groups used in the analysis are broad and may hide important differences among ethnic sub-groups. This is particularly important for the Asian group, among whom there is evidence of variation in Covid-19 outcomes between Bangladeshi, Pakistani and Indian groups (3, 6). Broad ethnic groups were analysed instead of more detailed groups to reduce bias caused by differences in the recording of ethnicity between the denominator data for populations (Census data) and information obtained on deaths using hospital activity data (HES data). The number of excess deaths among ‘Other’ ethnic groups should also be interpreted with some caution due to this potential mismatch.

Accounting for deprivation using quintiles of small areas does not identify deprived individuals or households within areas, so there may be greater effects of deprivation on excess deaths than identified in this study. Finally, the relative impact of the pandemic on sub-groups of the population may have varied throughout the pandemic period; by presenting the cumulative estimates we are not able to identify this variation.

Our findings need further investigation as to why these differences in mortality between ethnic groups independent of deprivation exist; a first step could be to look at underlying morbidities and aspects of deprivation not measured in the index used. Further work is also required to understand how the relationship between deprivation, ethnicity and excess mortality may have changed over the course of the Covid-19 pandemic and will change into the future. It is also important to examine where the deaths occurred as we know that during the pandemic to date there have been many excess deaths from causes other than Covid-19 that have occurred at home (12). What is not known is whether this was people choosing to die at home instead of hospital or people avoiding hospitals and experiencing an earlier death as a result. The relationship between excess deaths not directly attributable to Covid-19 and the time of death during the pandemic should also be explored. It is not clear the extent to which these are influenced by measures to control the spread of the virus, mortality displacement or longer-term implications of Covid-19 infection.

Our study shows that, similar to major historical pandemics in Europe, the Covid-19 pandemic has disproportionately impacted the most vulnerable and the most deprived (22-24). Effects of this sort are not limited to pandemics; the slowdown in improvement in life expectancy in England in the decade before the pandemic also disproportionately affected deprived populations and inequalities widened, but the pace of change was slow in comparison to that seen over the period examined in this study (25).

We illustrate the importance of examining the relationship between deprivation and excess deaths among defined age groups and ethnic groups. To date, routine surveillance of excess deaths during the Covid-19 pandemic both by ONS and PHE has not revealed this relationship; both organisations have reported results separately by age, deprivation and ethnic group (12, 26). Surveillance that reveals these inequalities will become increasingly important as the medium and long-term implications of the pandemic are revealed (24).

Even though analysis of death certificate information does highlight inequalities in mortality involving Covid-19, analysis of all-cause excess deaths shows that the impact on inequalities is even greater: the indirect effect of the pandemic on deaths has disproportionately affected people in ethnic minority groups and the most deprived White group. The Covid-19 pandemic has already increased inequalities in premature mortality between social and ethnic groups beyond those that existed in 2015–2019, and we do not yet understand the long-term implications, including the impact of ‘long Covid-19’ on mortality.

## Data Availability

Data are available upon reasonable request. All data relevant to the study are included in the article or uploaded as supplemental information.

## Contributors

SB, PF, SF, JF, AB, PB and PG contributed to conception of the study, study design and overall analysis plan. Specifically, in addition, SB, PF, JF, JN, YD and PG drafted the manuscript and revised the final paper; SF carried out the analysis and produced the graphs. All authors critically reviewed the final paper.

## Guarantor: SB

*SB affirms that the manuscript is an honest, accurate, and transparent account of the study being reported; that no important aspects of the study have been omitted; and that any discrepancies from the study as planned have been explained*.

## Funding

SB acknowledges funding received by the Australian Government through the Australian Research Council’s Centre of Excellence for Children and Families over the Life Course (Project ID CE200100025).

## Competing interests

None declared.

## Patient consent for publication

Not required.

## Patient and Public Involvement

It was not appropriate or possible to involve patients or the public in the design, or conduct, or reporting, or dissemination plans of our research

## Ethics approval

This study was carried out as part of the PHE responsibility to manage the COVID-19 pandemic. PHE has legal permission, provided by Regulation 3 of The Health Service (Control of Patient Information) Regulations 2002 to process confidential patient information (http://www.legislation.gov.uk/uksi/2002/1438/regulation/3/made) under Sections 3(i) (a) to (c), 3(i)(d) (i) and (ii) and 3(3) as part of its outbreak response activities. As such this work falls outside the remit for ethical review.

The study was subject to an internal review by the PHE Research Ethics and Governance Group and was found to be fully compliant with all regulatory requirements. As a full ethical review is not a requirement for this type of study and as no ethical or regulatory issues had been identified the study was approved.

## Copyright information

© Crown Copyright

---

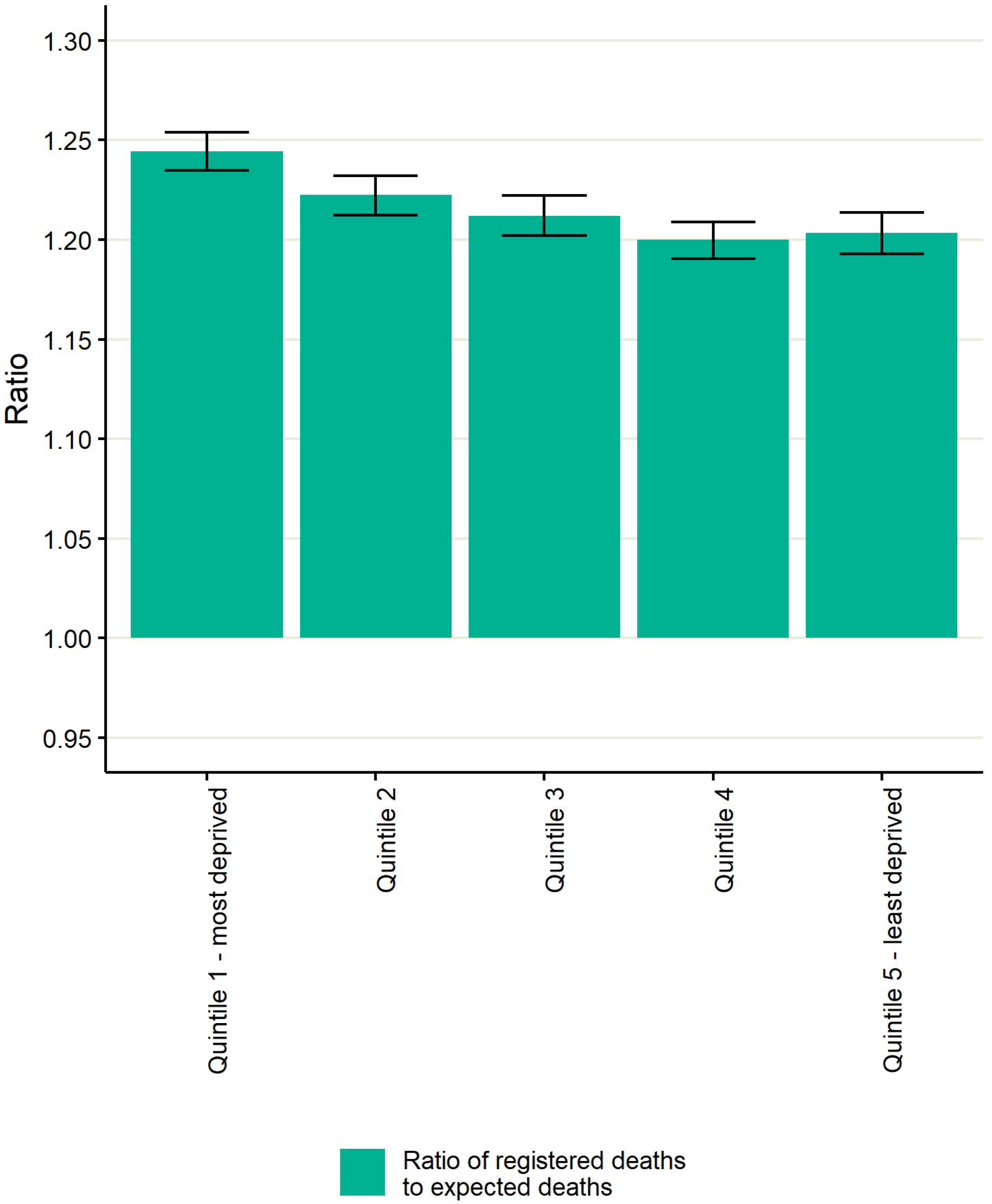

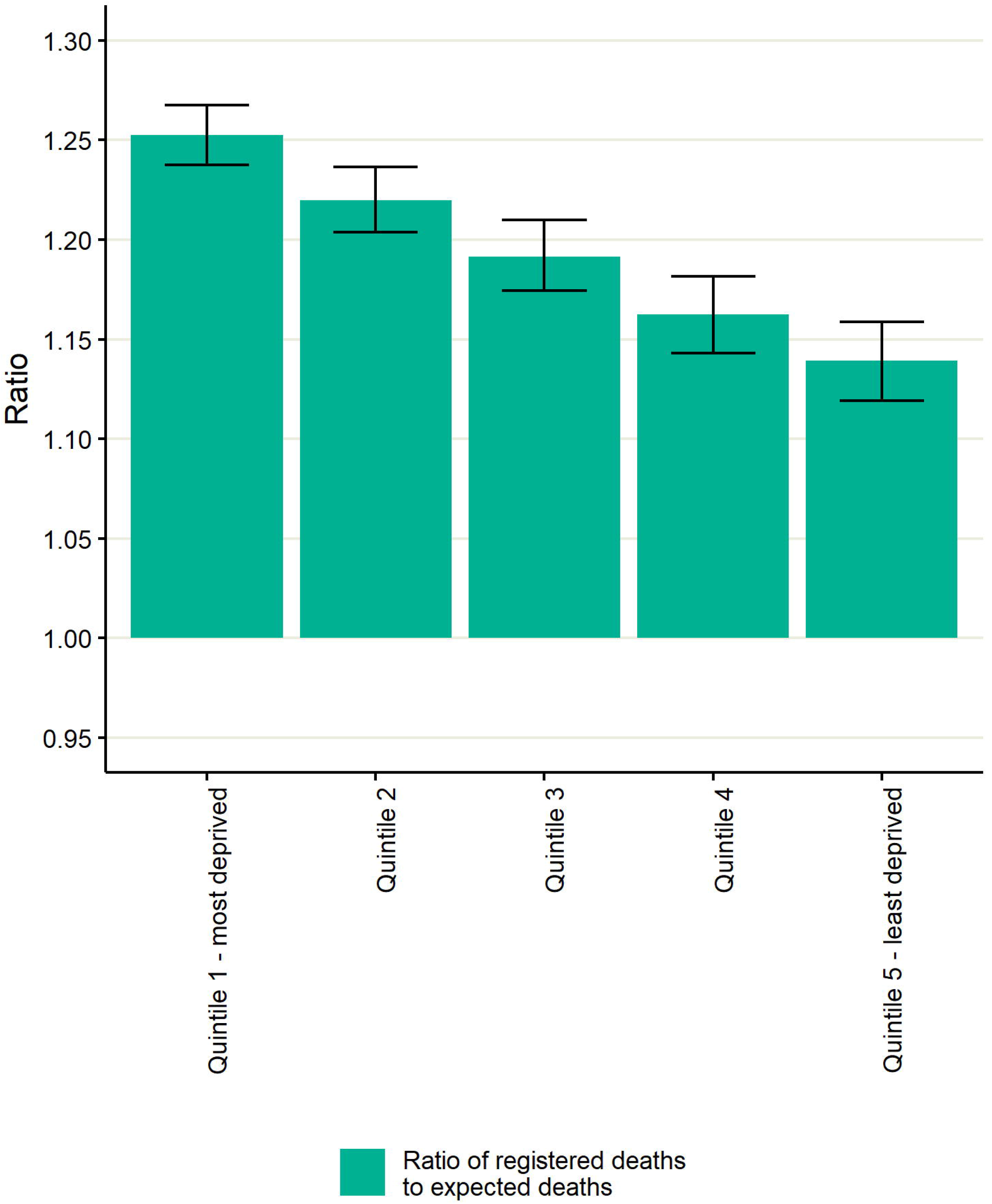

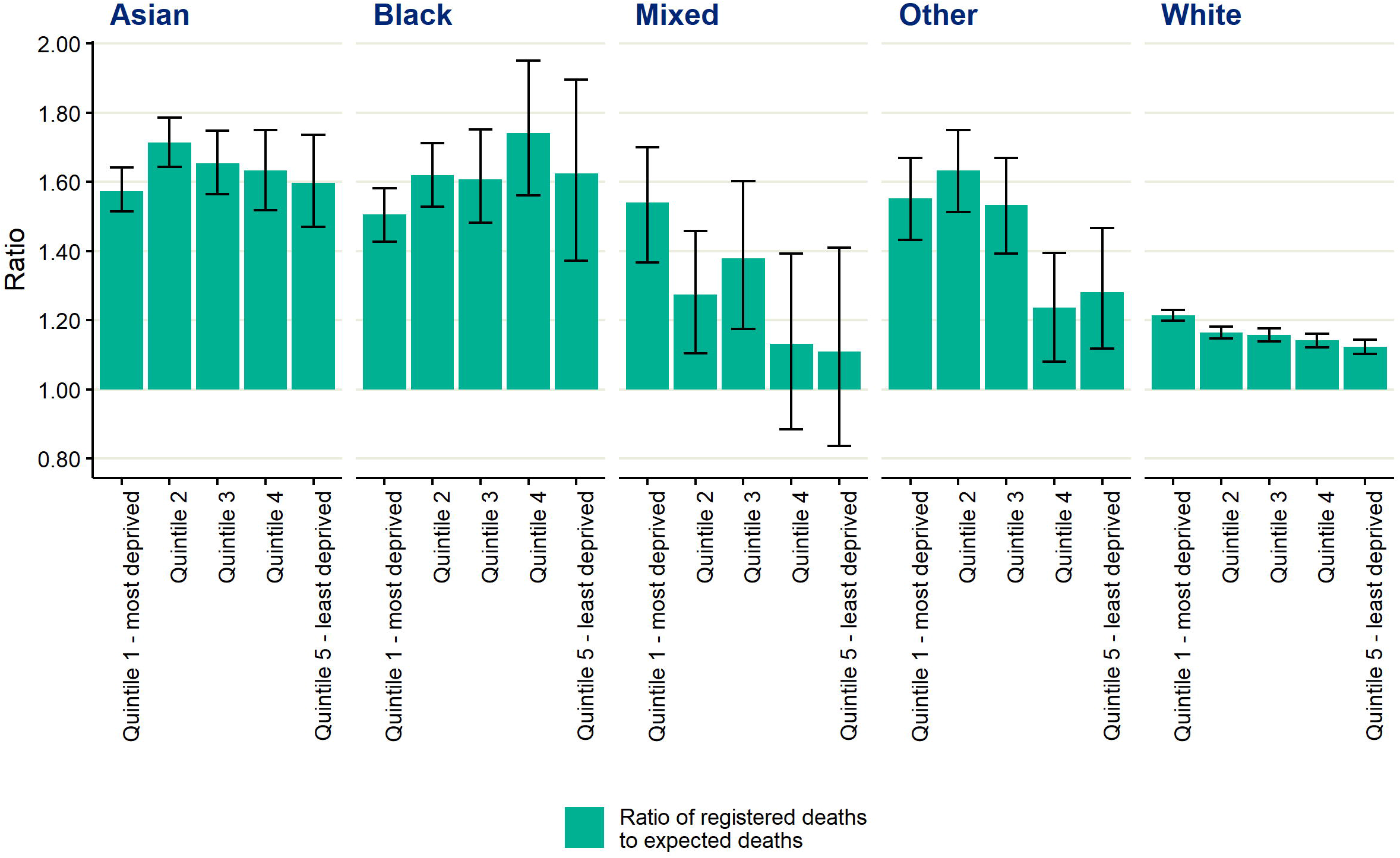

## Notes

### Competing Interest Statement

The authors have declared no competing interest.

### Funding Statement

Sharmani Barnard acknowledges funding received by the Australian Government through the Australian Research Council's Centre of Excellence for Children and Families over the Life Course (Project ID CE200100025).

### Author Declarations

This study was carried out as part of the PHE responsibility to manage the COVID-19 pandemic. PHE has legal permission, provided by Regulation 3 of The Health Service (Control of Patient Information) Regulations 2002 to process confidential patient information (http://www.legislation.gov.uk/uksi/2002/1438/regulation/3/made) under Sections 3(i) (a) to (c), 3(i)(d) (i) and (ii) and 3(3) as part of its outbreak response activities. As such this work falls outside the remit for ethical review. The study was subject to an internal review by the PHE Research Ethics and Governance Group and was found to be fully compliant with all regulatory requirements. As a full ethical review is not a requirement for this type of study and as no ethical or regulatory issues had been identified the study was approved.

